# Development and application of a Phytochemical Food Database (PhytoFooD) to assess the intake of dietary plant bioactives

**DOI:** 10.64898/2026.03.10.26348068

**Authors:** Costanza Michelini, Alice Rosi, Federica Bergamo, Cristiana Mignogna, Francesca Scazzina, Daniele Del Rio, Pedro Mena

**Affiliations:** Human Nutrition Unit, Department of Food and Drug, University of Parma, Parma, Italy

**Keywords:** Database, phytochemicals, food, diet, polyphenols, carotenoids, glucosinolates

## Abstract

Plant-based foods are complex systems, where a multitude of bioactive molecules, such as (poly)phenols and carotenoids are the outcome of endless interactions defining food chemical composition. Significant progress has been made to develop reliable food composition databases that can be used to assess the intake of dietary plant bioactives. However, many lesser-known phytochemicals, like glucosinolates and monoterpenoids are often excluded, also due to the fragmented information available in the literature. Therefore, we present PhytoFooD, a comprehensive phytochemical food database that collects qualitative and quantitative information on 1,067 bioactive compounds in 1,410 plant-based foods. We evaluated the intake of main plant bioactives in European diets and demonstrated the role of concentration variability within foods in intake assessments. This database represents a promising tool for dietary intake assessors and researchers in nutrition, paving the way for a comprehensive and accurate knowledge of our diet and the interconnected health effects of plant bioactives.

## 1. Introduction

Plant-based foods represent an extensive repository of important nutrients, such as macronutrients, vitamins, minerals, and a plethora of known and unknown putatively bioactive molecules, the health effects of which have yet to be fully understood (Menichetti et al., 2025). (Poly)phenols and carotenoids have been extensively investigated for their preventive effects against cardiovascular diseases and cancer (Liu, 2013), but many other plant bioactives may play a crucial role in human health. In contrast, excessive intakes of specific compounds such as caffeine and glycyrrhizin from coffee and liquorice, respectively, could lead to safety concerns (Deutch et al., 2019; Heckman et al., 2010). However, information on the content of these phytochemicals in national food composition databases is quite rare, and a scattered scenario is manifesting in the scientific literature. The incomplete chemical information of food composition greatly limits our knowledge on which set of compounds each individual is exposed to daily (also called ‘foodome’), and therefore, how dietary habits influence our health (Barabási et al., 2019). Analytical studies usually investigate single compounds or a few classes of phytochemicals in a limited number of foods, not allowing us to have a complete picture of the food composition. Our diet, though, is far from being composed of a few foods, supplying instead more than 139,000 chemicals (Menichetti et al., 2024). The absence of a comprehensive and reliable food composition database for plant bioactive compounds remains a challenge for the scientific community. Although important steps have been taken in the last twenty years to fill this gap, such as the development of the Periodic Table of Food Initiative (Jarvis et al., 2024) and the EUROFIR eBASIS database (Kiely et al., 2010), much information on the concentration of chemicals in plant-based foods is still missing. The elevated number of dietary sources, the multiple phytochemical classifications, the diversity of analytical methods, and the wide variability of compound concentrations within foods are just some of the major challenges of developing a food composition database (Scalbert et al., 2011). These issues undermine the reliability of the phytochemical intake assessment, leading, in some cases, to inconsistent or incomparable results (Lanuza et al., 2022).

In this context, our aim was to develop a Phytochemical Food composition Database (PhytoFooD) that gathers concentration data of all dietary plant bioactive compounds in a single and unique databank. To demonstrate its potential applications, we investigated the intake of major phytochemicals from a subset of representative foods that are among the most consumed by the European population. Finally, we examined how the concentration variability of chemicals within foods may affect intake assessment results, providing insights into caffeine intake from espresso coffee in the European population and at the individual level in an Italian cohort, as a paradigmatic case.

## 2. Materials and methods

### 2.1 Development of PhytoFooD

Dietary plant bioactives have been hierarchically classified into 4 main families, according to PhytoHub classification (https://phytohub.eu/) and some structural readjustments. Briefly, they have been categorized into (poly)phenols, terpenoids, *N*-containing compounds, and miscellaneous phytochemicals. Each family includes multiple classes and subclasses (Table S1). Afterward, a methodological procedure consisting of two main steps was implemented to create a comprehensive Phytochemical Food Database, which gathers composition data on bioactive compounds in plant-based foods from national food composition databases and the most up-to-date scientific literature.

#### 2.1.1 Data collection

Information on the content of plant bioactives in food commodities have been firstly retrieved from 13 food composition databases, namely: Phenol-Explorer (Neveu et al., 2010), eBASIS (Kiely et al., 2010), USDA FoodData Central databases (Foundation Foods, SR Legacy and FNDDS) (https://fdc.nal.usda.gov/), Dr. Duke’s Phytochemical and Ethnobotanical Databases (https://phytochem.nal.usda.gov/), Global Food Composition Database for Phytate (FAO/IZiNCG, 2018. FAO/INFOODS/IZiNCG Global Food Composition Database for Phytate Version 1.0 - PhyFoodComp 1.0. Rome, Italy), Ciqual (https://ciqual.anses.fr/), Frida Food Data (https://frida.fooddata.dk/), Fineli (https://fineli.fi/), Standards Tables of Food Composition in Japan (https://www.mext.go.jp/en/policy/science_technology/policy), FOODfiles (https://www.foodcomposition.co.nz/), and, finally, CoFID (https://www.gov.uk/government/publications/composition-of-foods-integrated-dataset-cofid).

Additionally, an extensive literature review in Scopus (www.scopus.com) was conducted to enhance the reliability of the information collected, particularly for those classes where data from food composition databases were limited. The bibliographic search of peer-reviewed original articles and reviews, which started in November 2022 and concluded in March 2025, was fragmented into sub-revisions specifically developed for the classes and subclasses of compounds. Combinations of words related to four main topics, being ‘phytochemicals’, ‘foods’, ‘concentration’, and ‘diet’ were used. Inclusion and exclusion criteria regarding the type of study, the analytical method used, and the year and journal of publication were established to ensure high data quality, with thresholds varying among compounds depending on the evidence available. Particularly, the most recent analytical studies published in peer-reviewed journals have been considered; the use of a high-resolution analytical techniques, such as liquid/gas chromatography coupled to mass spectrometry, was a key criterion for our literature search. Specifics, both related to the article and food analysed, have been firstly reported in separate spreadsheet files -renamed ‘archives’- where concentration data were harmonized in mg/100 g of food item. Finally, each food and related chemical data from databases and/or scientific publications were matched univocally with our reference database, based on the Food Composition Database for Epidemiological Studies in Italy BDA (https://bda.ieo.it/). This match was primarily based on the similarities in food names and macronutrient composition between foods. If not already found in the BDA database, new food items were created and added to the final PhytoFooD food list. When available, concentration data in processed foods have been also collected. Alternatively, yield factors obtained from CREA (Lisciani et al., 2022) and Bognar’s tables (Bognár, n.d.), were used for food processing.

#### 2.1.2 Data analysis

This step provided valuable information on the chemical concentrations in foods that are usually missing in food composition databases. The data collection yielded a highly variable amount of information for each compound and food, which required prior hand-curated processing before obtaining the final records. Therefore, for each dataset of bioactive concentrations in foods, central and deviation measures were calculated, including the average concentration, the standard deviation, the minimum, the maximum, and the median concentrations, and the coefficient of variation. The application of criteria, when performing the literature search, allowed us to select the most relevant and reliable data, but the wide variability in chemical concentrations in foods remained a significant challenge. Hence, with a dataset size ≥ 3 for each food-compound binomial, multiple tests were applied to exclude outliers:

- The first test consisted of measuring the level of dispersion of the value considered to the central point when applying two standard deviations (mean ± 2 SD).
- The second test comprised the Grubb’s test (Grubbs, 1969). An outlier is identified if the calculated Z score, expressed as (mean – value)/SD, exceeds a predetermined threshold value that depends on the dataset size.
- The third and last test consisted of Tukey’s test (Hoaglin et al., 1986). It detects the presence of an outlier if the concentration value is lower/higher than 1.5 times the interquartile range (IQR) from the first/third quartile (Q1, Q3) (i.e. x < Q1 − 1.5 IQR, x > Q3 + 1.5 IQR).

Two out of three positive results have been considered sufficient to define the value as an outlier and to exclude it from the dataset. Consequently, central and deviation measures were recalculated without the detected outlier(s). Finally, a robustness index was calculated for the obtained results for each food-compound combination, based on the dataset size considered. A robustness index from 1 to 3 has been applied to datasets (excluding outliers) with n < 3, 3 ≥ n < 5, and n ≥ 5, respectively.

A slightly different approach was used in the development of (poly)phenol, monoterpenoid, and thiosulfinate tables, where two specialised food composition databases containing wide information -namely Phenol-Explorer and Dr. Duke’s Phytochemical and Ethnobotanical Databases- were the principal data sources. In the case of (poly)phenols, additional data has been added (Rosi et al., 2021; Ziauddeen et al., 2019) to fill major gaps currently present on Phenol-Explorer and precluding a comprehensive dietary assessment.

### 2.2 Food consumption data

#### 2.2.1 European cohorts from the EFSA database

Dietary information of European adults (18-65 years old) was extracted from the most recent national surveys, collected in the EFSA Comprehensive European Food Consumption Database (https://www.efsa.europa.eu/en/data-report/food-consumption-data) and harmonized using the FoodEx2 classification. Food consumption data from 22 European countries, being Belgium, Bosnia and Herzegovina, Croatia, Cyprus, Czechia, Estonia, Finland, France, Germany, Greece, Hungary, Ireland, Italy, Latvia, Netherlands, Poland, Portugal, Romania, Serbia, Slovenia, Spain and Sweden were used. To reach the utmost level of completeness and reflectiveness of European dietary habits, different grades of specificity – or Exposure hierarchy levels – have been considered to rank the most consumed food items by the European population. Finally, daily consumption data on 20 representative foods/food categories, among the most consumed, have been selected.

#### 2.2.2 Oral (Poly)phenol Challenge Test Cohort (OPCT)

Food consumption data of 297 healthy volunteers (19-73 years old) enrolled in the OPCT study (Mena et al., 2022) were also considered in our assessment. The trial was approved by Ethics Committee of the Area Vasta Emilia Nord region (Italy, 1352/2020/SPER/UNIPR OPCT), registered (ClinicalTrials.gov, NCT05414084) and participants provided informed consent. During this acute intervention study, participants were asked to provide information on their dietary habits using the EPIC food frequency questionnaire (Pisani, 1997). Daily consumption data of espresso coffee was used to estimate caffeine intake, providing insights into phytochemical intake at the individual level.

### 2.3 Association between PhytoFooD and dietary intake data

Food items with the highest level of specificity (Exposure hierarchy L7 of FoodEx2 classification system) from the EFSA database were matched with our PhytoFooD food list, based on food names and macronutrient concentrations; alternatively, the most similar food from our catalogue was used. This was done for every country data. Major dietary phytochemicals found in the selected foods were considered to provide insights into the plant bioactive intakes in the European population (Table 1 a-b). Daily intakes of phytochemicals (mg/day) were estimated using the mean food consumption data (grams/day) and the average concentration of chemicals in the same foods (mg/100g of food). When required, food-specific yield factors were applied to retrieve missing information about the content of phytochemicals in treated foods. The same approach was applied to the OPCT dietary data. Briefly, espresso coffee consumption data was matched with the corresponding food item from the PhytoFooD, and different concentrations of caffeine (the minimum, the average, and the maximum) were applied to investigate the extent of its intake variability. The food match and the phytochemical intake assessment were performed using Microsoft Excel and Access softwares. Graphical representations have been created using R version 4.5.1 software (R Core Team 2025) (Makowski et al., 2021) (https://www.r-project.org/) and RStudio version 418 (Posit Software, 2025) (https://posit.co/), particularly using the following packages: dplyr (Wickham, François, et al., 2014), ggplot2 (Wickham, 2016), rnaturalearth (Massicotte & South, 2017), rnaturalearthdata (South et al., 2017), sf (Pebesma, 2018; Pebesma & Bivand, 2023), and tidyr (Wickham, Vaughan, et al., 2014).

**Table 1:**
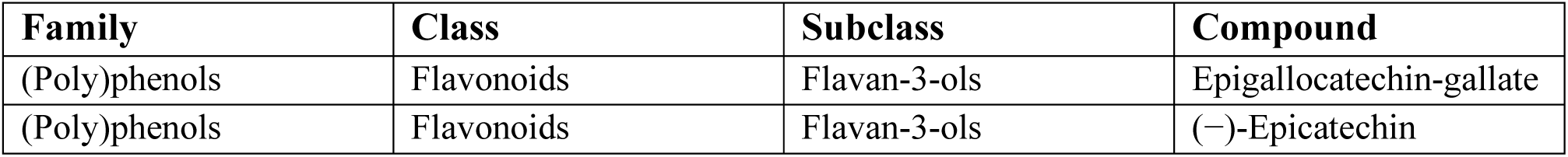
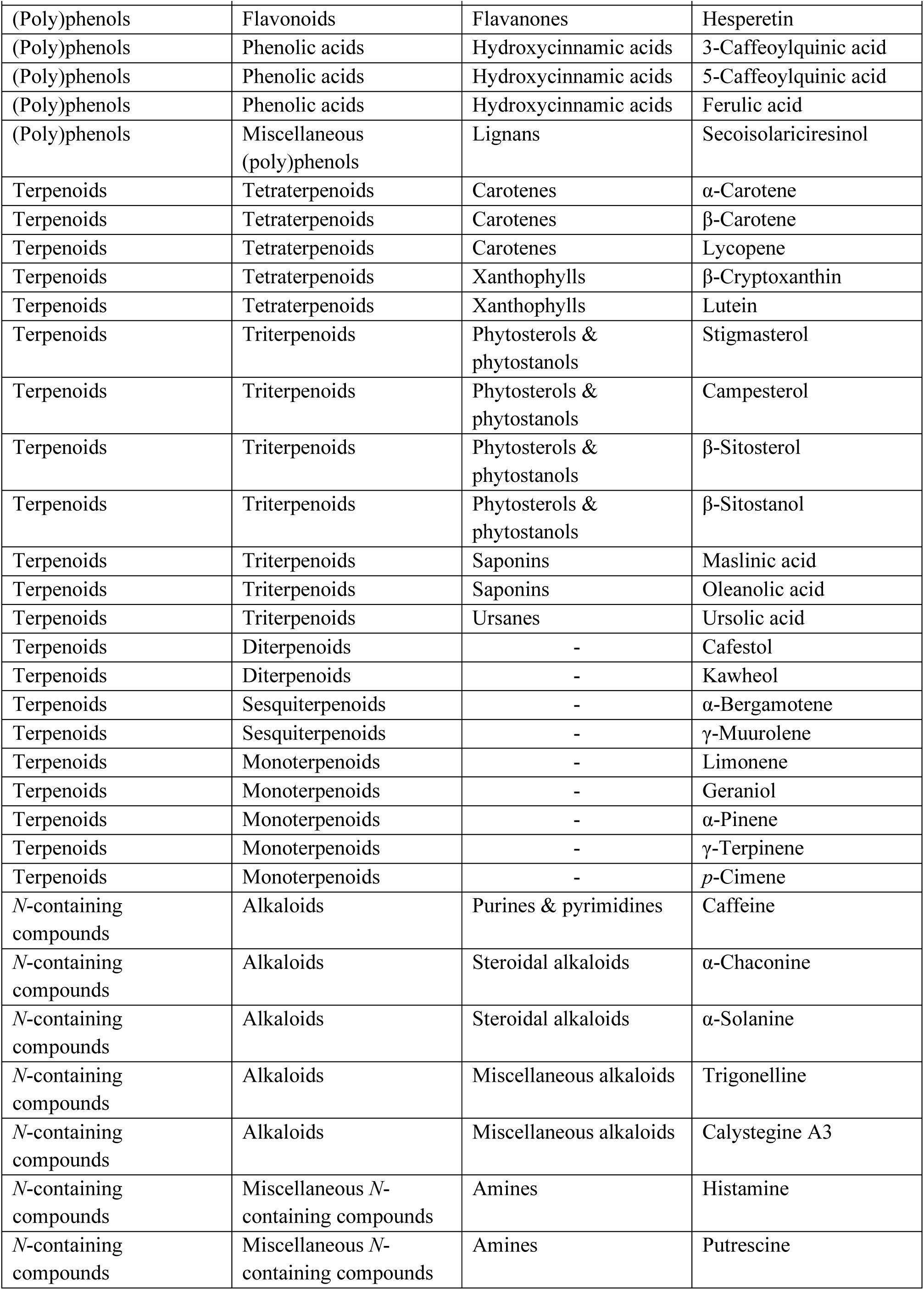
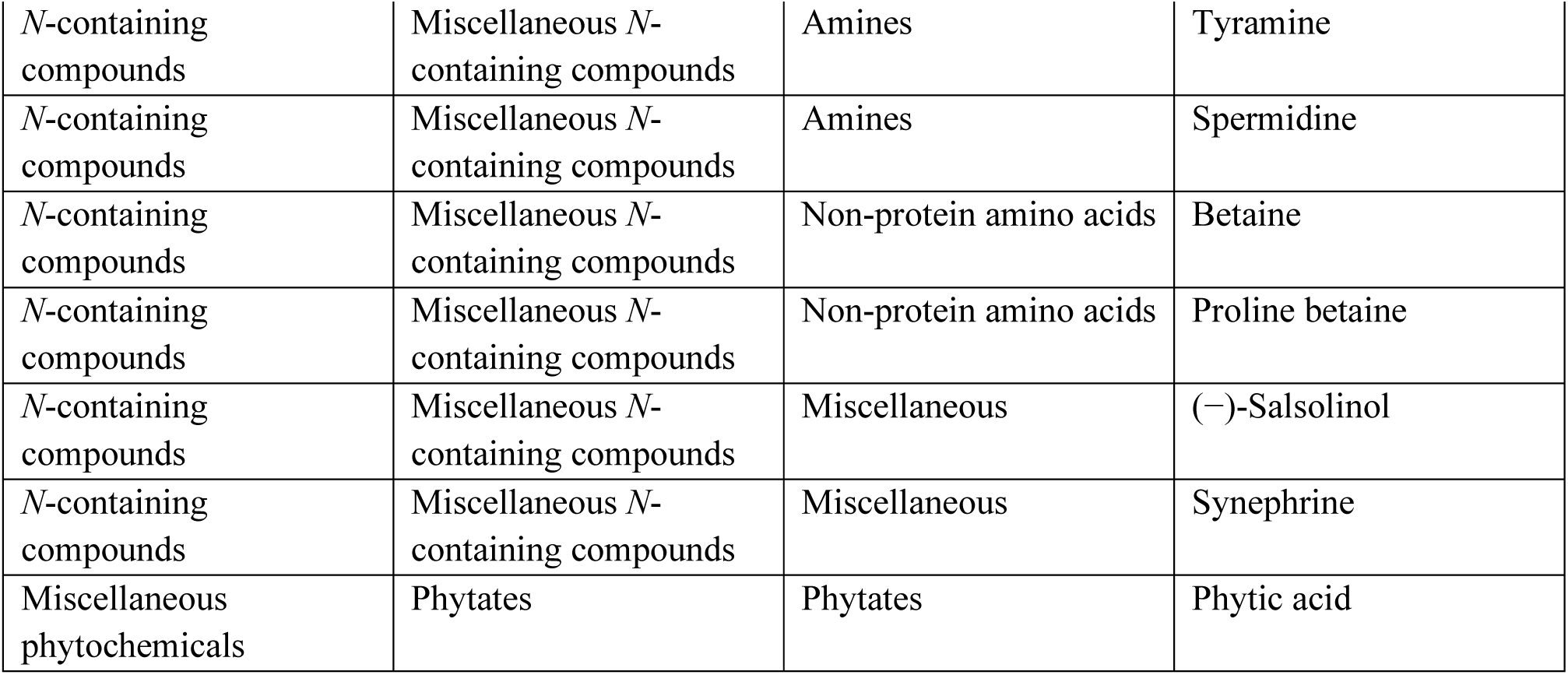
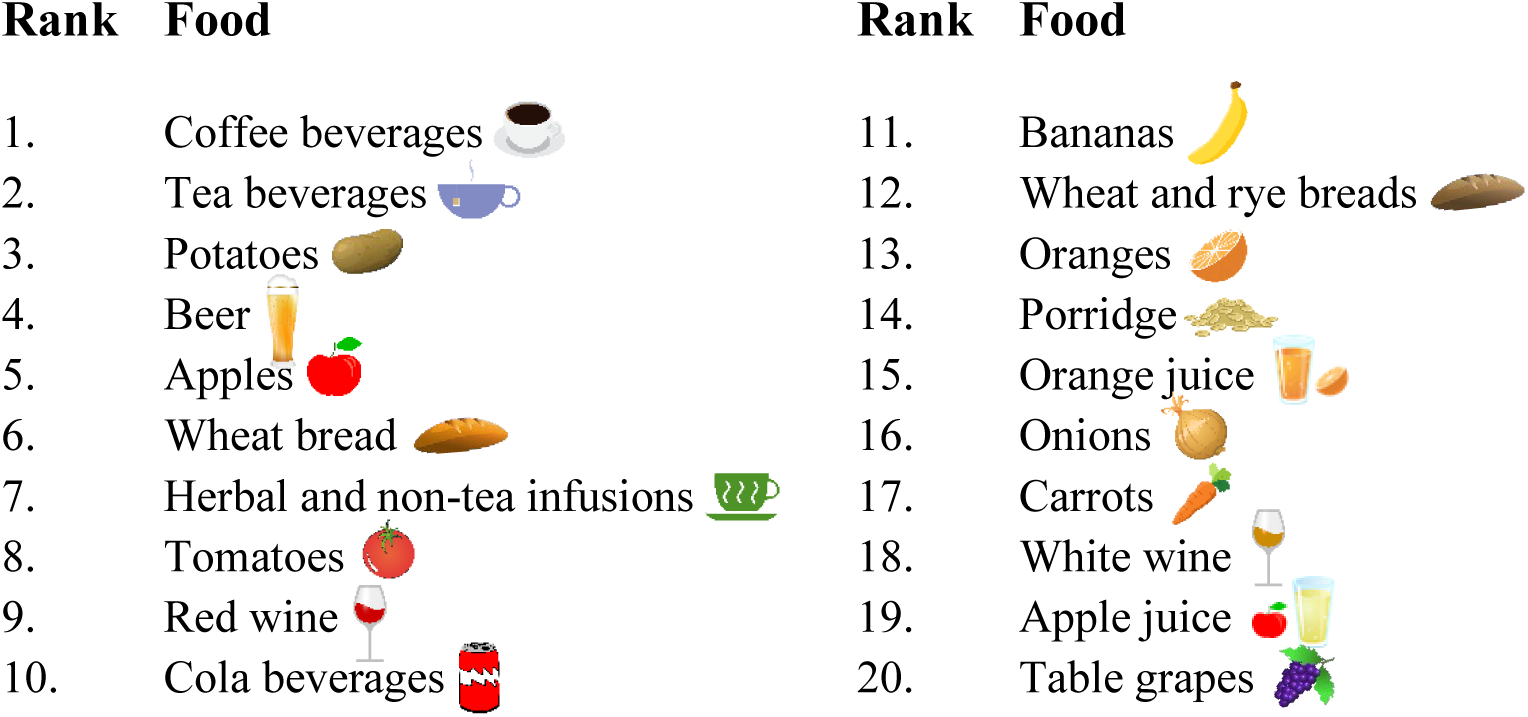
Major dietary phytochemicals found in most consumed foods by the European population. We selected the most abundant dietary phytochemicals (a) present in the 20 most consumed foods (b) among European countries, to demonstrate a potential application of PhytoFooD in a phytochemical intake assessment context.

## 3. Results

### 3.1 The PhytoFooD properties

Currently, the Phytochemical Food Database contains 41,130 mean concentration values regarding 1,067 phytochemicals in 1,410 foods, the latter belonging to 15 food categories (Figure 1). Information on the quantity of phytochemicals in foods has been compiled from 13 food composition databases and over 330 scientific original articles. The PhytoFooD database consists of multiple tables recorded in Excel workbooks, each of them summarized in ‘overview tables’ showing: the food item names, the complete set of central and deviation measures of the bioactive compounds, and, lastly, related robustness indexes (Table 2). As a result, each food has been extensively profiled for its phytochemical content. For instance, Figure 2 shows the list of plant bioactives contained in espresso coffee, based on the information collected. Along with a deep chemical characterization, details on the concentration variability within foods have been provided, when sufficient data were available. A considerable fluctuation in bioactive concentrations has been observed for most phytochemicals in foods, as briefly shown in Table 2. This emphasises the complexity of foods and how influential factors such as the type of cultivars, environmental conditions, and seasonality can be on the chemical composition of foods. Ultimately, 17% of the final concentration panels showed the highest level of robustness (3), meaning that a minimum of five data points were collected to obtain those results. Nearly 15% presented a robustness index of 2 (3 ≥ n < 5), and, finally, ∼ 68 % displayed a level 1 (n < 3). These results did not consider (poly)phenol, monoterpenoid and thiosulfinate tables, since they have been developed using two highly-curated databases, namely Phenol-Explorer (Neveu et al., 2010) and Dr. Duke’s Phytochemical and Ethnobotanical Databases (https://phytochem.nal.usda.gov/) as main data sources.

**Figure 1:**
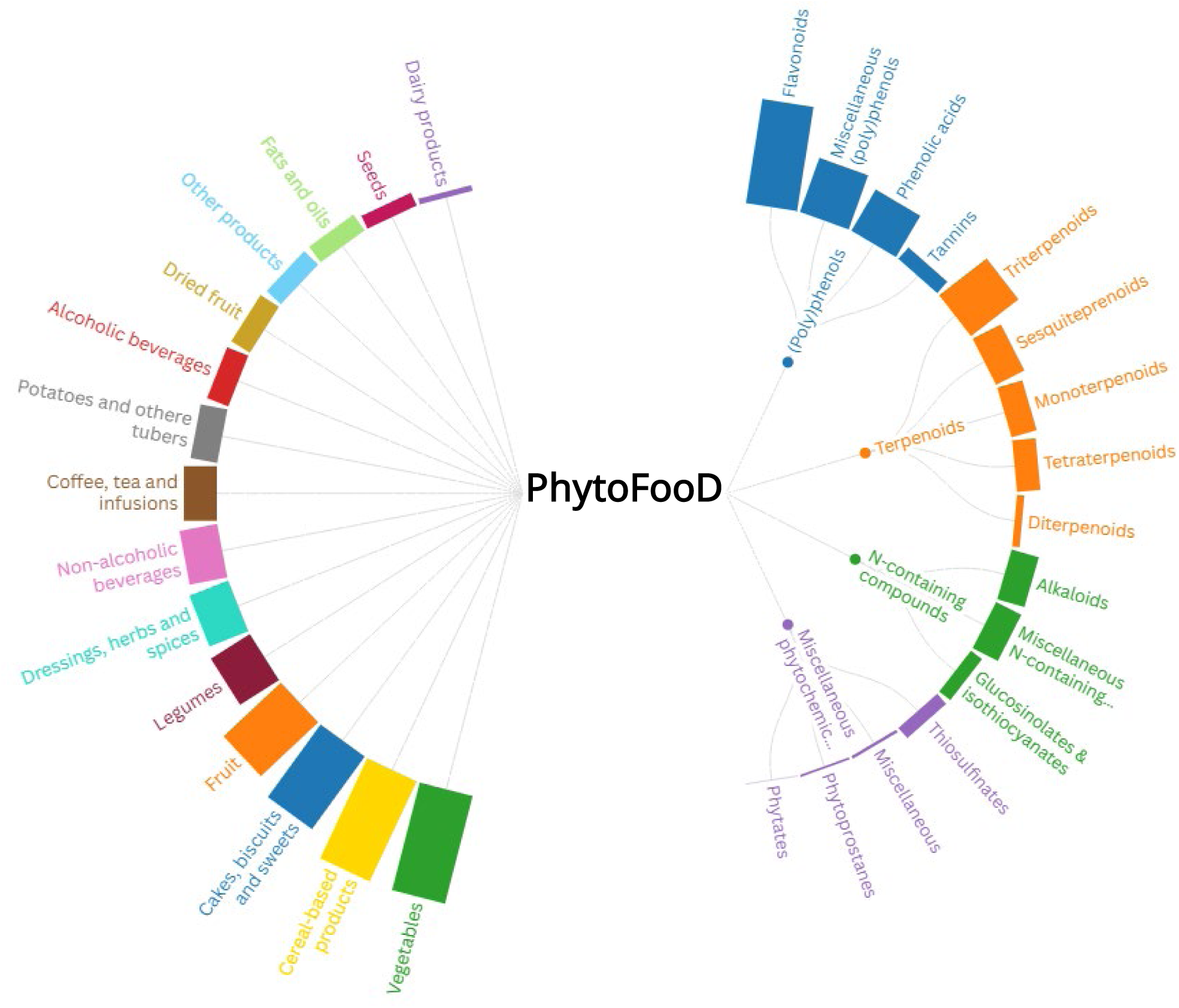
Classification of phytochemicals and foods in PhytoFooD. The Phytochemical Food Databases is a comprehensive databank that collects information about 1,067 bioactive compounds in 1,410 plant-based foods and composite foods containing plant-based ingredients. Bar length refers to the amount of data collected.

**Figure 2:**
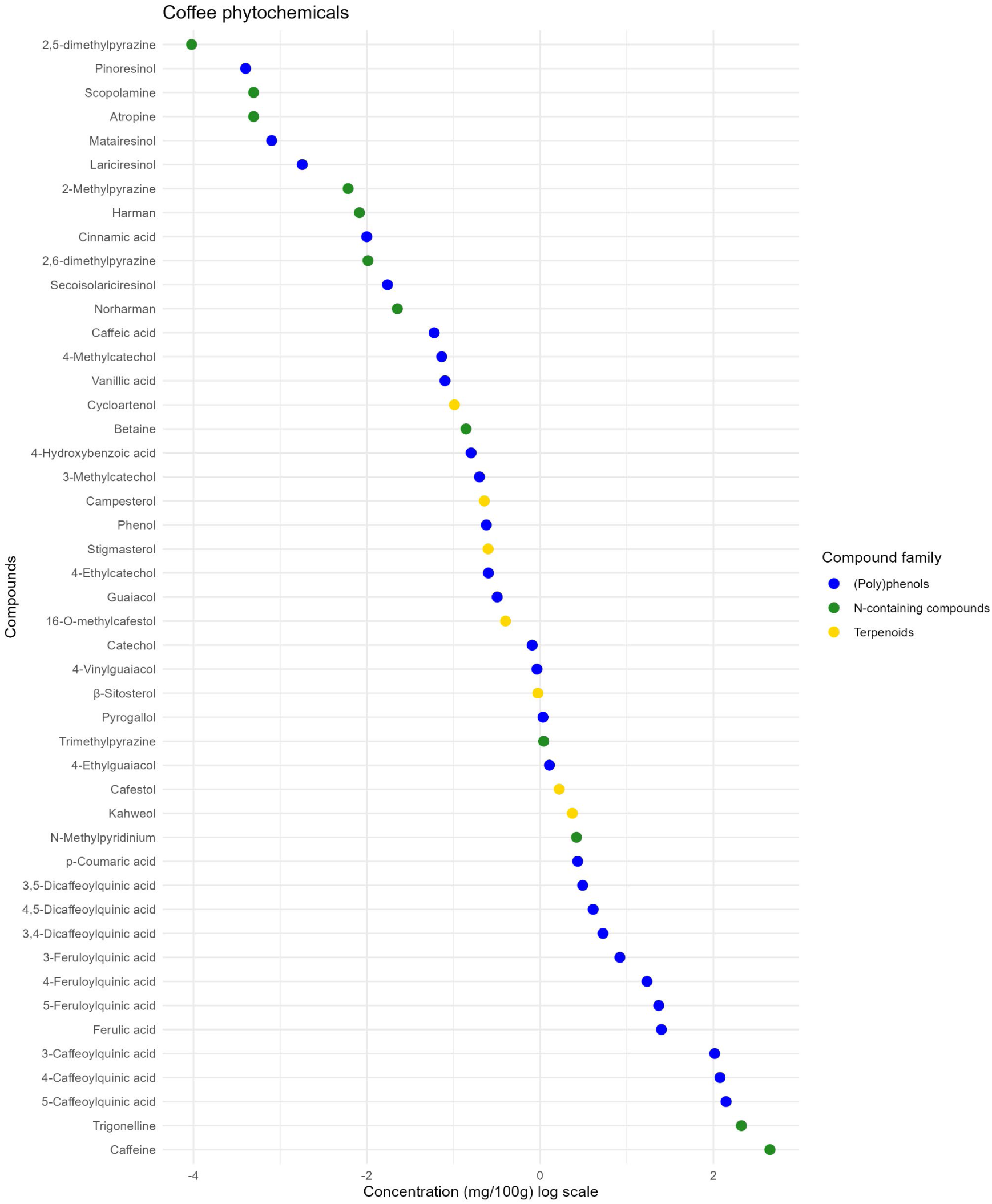
Chemical profile of the espresso coffee. Coffee represents one of the most important food sources of phytochemicals in the diet of the European population. It provides a wide range of bioactive molecules, belonging to different compound families and with broad concentration differences.

**Table 2:**
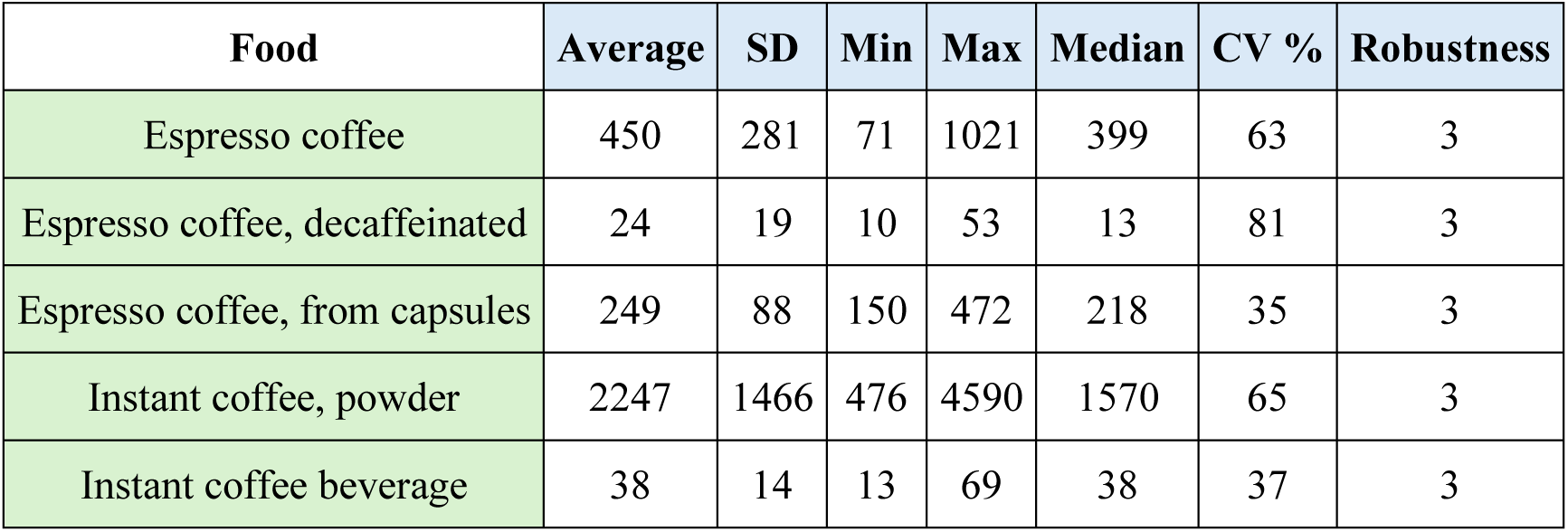
Brief visualization of PhytoFooD overview tables: the caffeine example. Summary tables of the PhytoFooD are provided for phytochemical classes and subclasses, with information about the food item name, the average concentration, the standard deviation, the minimum, the maximum and the median concentrations, the coefficient of variation (%) and, finally, the robustness index -based on data availability-.

| Food | Average | SD | Min | Max | Median | CV % | Robustness |
| --- | --- | --- | --- | --- | --- | --- | --- |
| Espresso coffee | 450 | 281 | 71 | 1021 | 399 | 63 | 3 |
| Espresso coffee, decaffeinated | 24 | 19 | 10 | 53 | 13 | 81 | 3 |
| Espresso coffee, from capsules | 249 | 88 | 150 | 472 | 218 | 35 | 3 |
| Instant coffee, powder | 2247 | 1466 | 476 | 4590 | 1570 | 65 | 3 |
| Instant coffee beverage | 38 | 14 | 13 | 69 | 38 | 37 | 3 |

### 3.2 Application of PhytoFooD within a European framework

Food composition databases are fundamental tools that provide insights into the nutritional intake of a population. To provide a practical example of the PhytoFooD potential uses in this context, we investigated the intake of major phytochemicals from the most consumed plant-based foods by the European adult population (Table 1). The most recent national surveys involving adults aged 18 to 65 years old were selected, resulting in 22 countries considered in our assessment. Figure 3 represents the total daily intakes of the phytochemicals investigated in this work in the European countries, gathered in (poly)phenols, terpenoids, *N*-containing compounds and miscellaneous phytochemicals. Detailed information on the contribution of most consumed food items to the intake of each compound family in various countries is presented in Figure 4. Netherlands, Finland, and Germany showed the highest intake of (poly)phenols, reaching ∼600 mg/day. Coffee was the primary food source in all countries. Among fruits and vegetables considered in this assessment, oranges and carrots were the principal sources of terpenoids. Portugal, Spain, and Greece proved to have the highest intake of these compounds, at ∼500 mg/day. Within the *N*-containing compound family, caffeine from coffee was the prevalent phytochemical consumed, frequently representing more than 50% of the total intake. This occured, for instance, in Serbia, Bosnia and Herzegovina, and Romania, which intakes of *N*-containing compounds ranged from 800 to 1200 mg/day, with a caffeine intake from ∼400 to ∼700 mg/day. Lastly, potatoes and wheat and bread rolls turned out to be the major food sources of miscellaneous phytochemicals, in this assessment fully represented by phytic acid. Romania, Czechia, and Hungary were the three European countries with the highest intake, around 240 mg/day. Detailed data on the intake of principal phytochemical classes and subclasses from the most consumed plant-based foods in European countries are shown in Figure S1, providing insights into the diversity of plant bioactive intake among populations.

**Figure 3:**
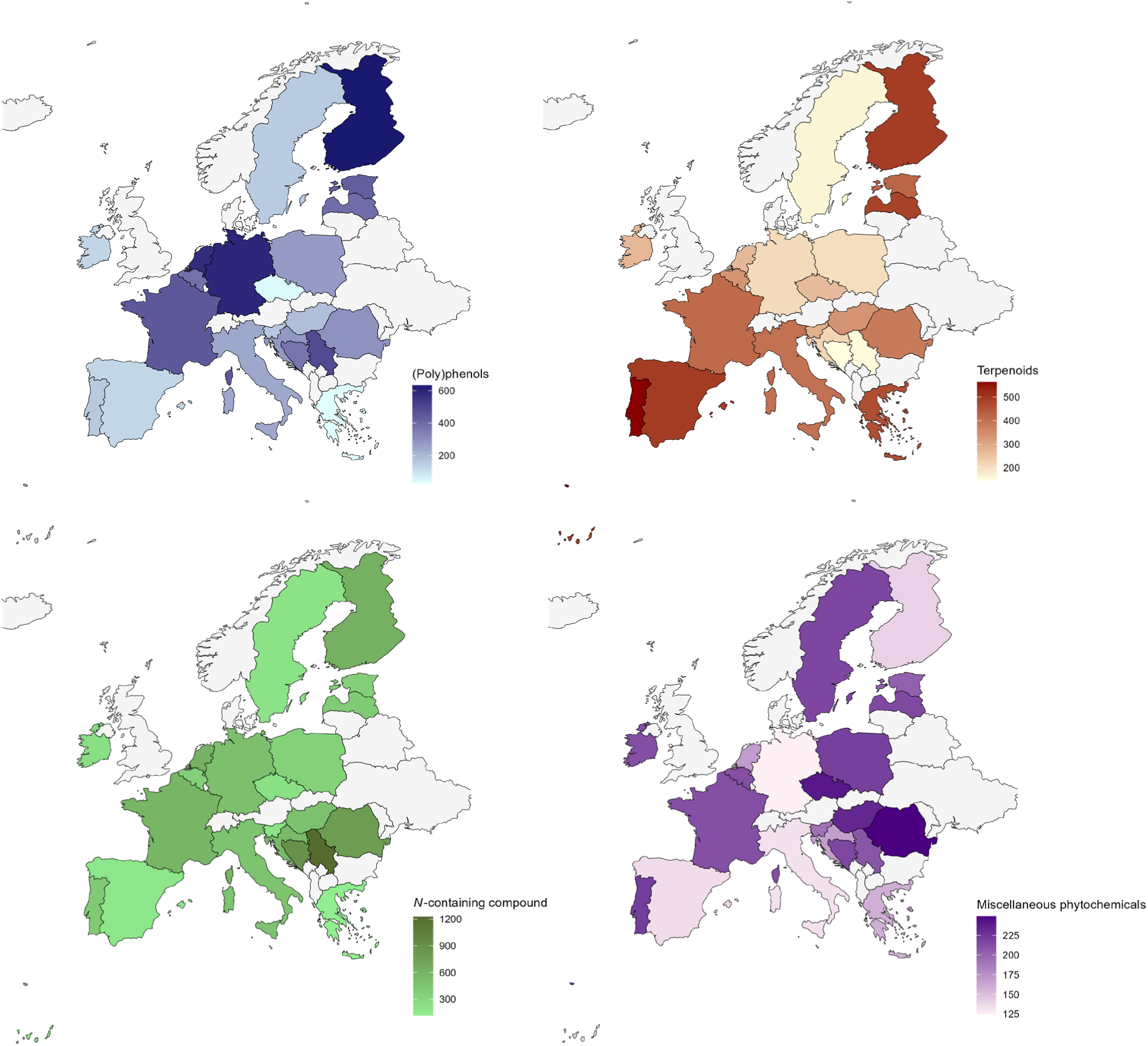
Geographic heatmap of Europe about dietary phytochemical intakes. We selected major plant bioactives found in 20 representative food items among the most consumed in the European population, to provide insights into the dietary phytochemical intakes. Blank spaces in the geographic heatmaps indicate countries of which phytochemical intake data are missing.

**Figure 4:**
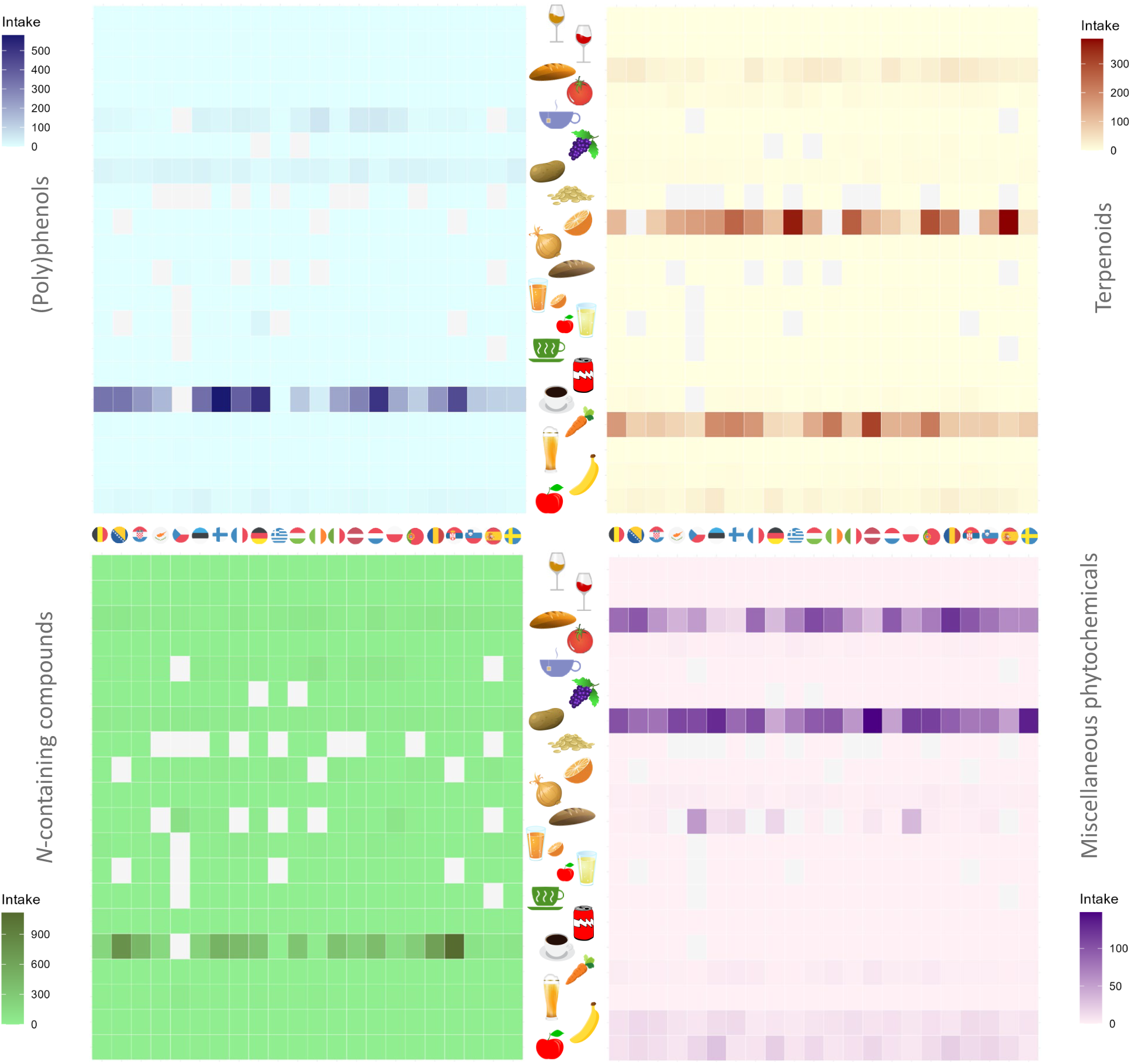
Contribution of food items to the total intake of phytochemical families in the European countries. Food items represented in the graph are reported in the following order: white wine, red wine, wheat breads, tomatoes, tea beverages, table grapes, potatoes, porridge, oranges, onions, wheat and rye breads, orange juice, apple juice, herbal and non-tea infusions, cola beverages, coffee beverages, carrots, beer, banana and apples. Flags correspond to this sequential order of European countries: Belgium, Bosnia and Herzegovina, Croatia, Cyprus, Czechia, Estonia, Finland, France, Germany, Greece, Hungary, Ireland, Italy, Latvia, Netherlands, Poland, Portugal, Romania, Serbia, Slovenia, Spain and Sweden. Blank spaces in the heatmaps denote that consumption data of specific foods are missing.

### 3.3 Application of PhytoFooD to phytochemical food concentration variability

Foods naturally exhibit variability in the concentration of phytochemicals due to factors related to their intrinsic characteristics and the surrounding environment. Unfortunately, this quantitative information is lacking in most food composition databases, where only average concentration data of chemicals in foods are usually reported. This Phytochemical Food Database has been created with the supplementary aim of providing insights into the influence of phytochemical food concentration variability on the dietary intakes of plant bioactives.

As an example with potential nutritional benefits and toxicological facts, coffee is one of the most consumed foods in Europe and an important source of bioactive molecules such as caffeine. However, its content is highly variable, even within the same type of coffee: for instance, a cup of espresso coffee (30 mL) might contain from 21 to 306 mg of caffeine, with an average concentration of 135 mg. Applying this range and values to coffee consumption data yields a broad spectrum of intake values. Figure 5 represents this variety at both the population (Fig. 5a) and individual levels (Fig. 5b), using average consumption data of espresso coffee from 14 European countries and from 297 Italian volunteers from the Oral (Poly)phenol Challenge Test -OPCT- study. Particularly, when the highest concentration of caffeine in espresso coffee is used, occasional exceeding of the 400 mg/day safe threshold -established by EFSA (EFSA Panel on Dietetic Products, Nutrition and Allergies (NDA), 2015)- could be observed at the population level. This trend is increasingly evident and dominant when dietary data of individuals are used. In the case of the OPCT participants, the safe threshold is highly exceeded when both average and maximum concentrations of caffeine are considered, with the gap getting wider as the average amount consumed by each individual increases.

**Figure 5:**
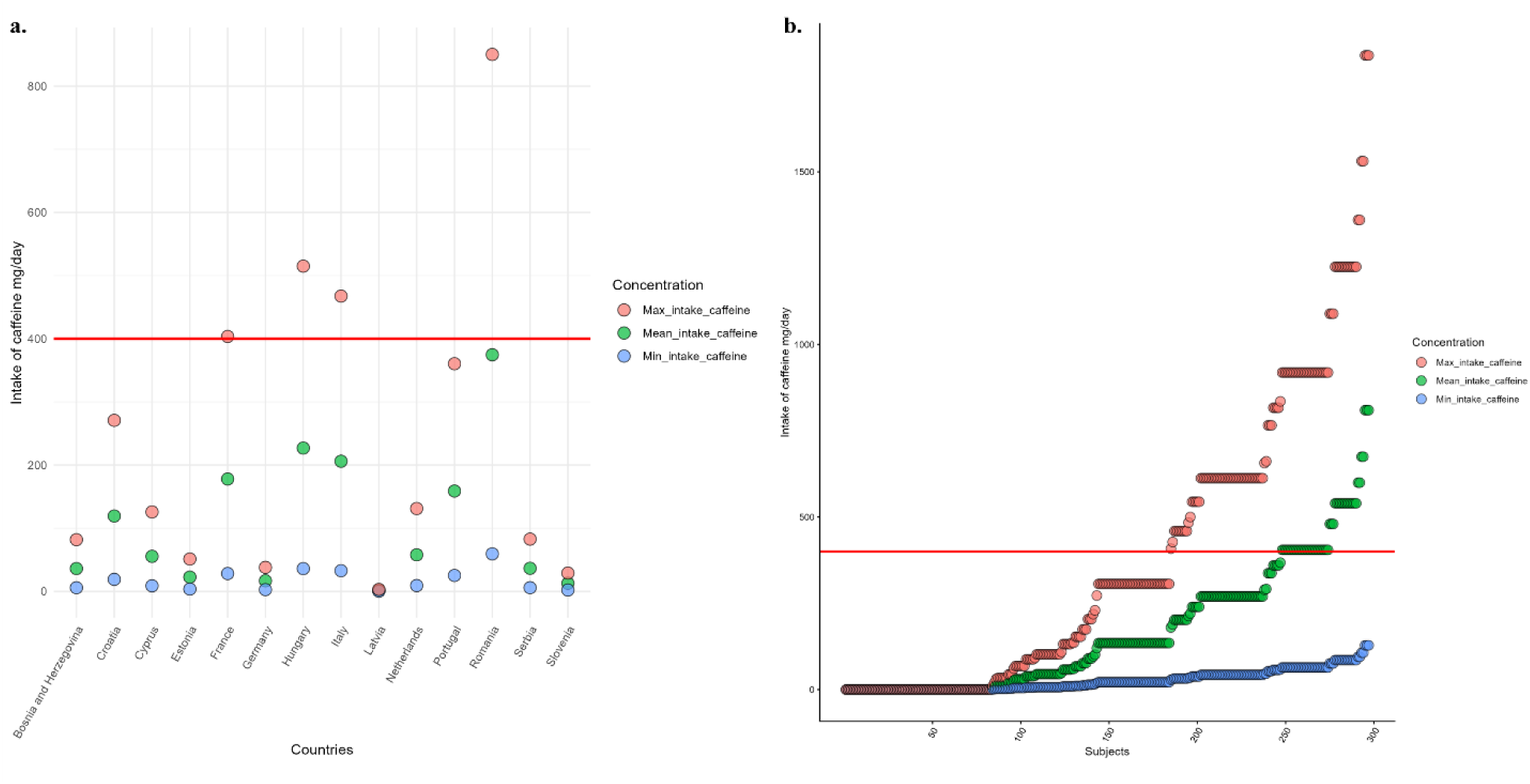
Effect of caffeine concentration variability in espresso coffee on its dietary intake: application at population and individual level. Caffeine intake from espresso coffee was estimated in European countries (a) and volunteers of the OPCT study (b), using the minimum, the average and the maximum concentration of caffeine reported in an espresso coffee in our database. The red line at 400 mg/day is the threshold indicated as safe for adults by EFSA.

## 4. Discussion

PhytoFooD compiles information on the content of over 1000 bioactive compounds in a diverse range of foods, creating a unique databank that yields the most comprehensive phytochemical food composition database to date. It collects data about well-known bioactive compounds, such as (poly)phenols and carotenoids, and many more chemicals poorly considered, like low-molecular-weight terpenoids, alkaloids, and glucosinolates. The curated data collection from the most relevant and up-to-date literature, as well as national food composition databases from around the world, enabled us to create an instrument that can be used globally, with numerous potential applications. Each food is extensively profiled from a chemical perspective, based on the results of our implemented data collection and analysis procedure. When available, information on the phytochemical concentration variability within foods is provided. Lastly, a robustness index value has been associated with the obtained results, furnishing additional information on the data quality of each food and compound. These features provide key evidence that most food composition databases overlook.

Despite these valuable qualities, some limitations must be acknowledged: firstly, the shortage of concentration data of lesser-known phytochemicals in foods, such as low-molecular-weight terpenoids, alkaloids, and glucosinolates, among others, leads to an incomplete food characterization and consequent dietary assessment. This could be due to the structural complexity of plant bioactives and food matrix interactions that do not always allow researchers to properly characterise all phytochemicals in foods (Parada & Aguilera, 2007). Although significant technological advances have been made in recent years, the multitude of analytical methods and techniques used to identify and quantify phytochemicals in foods may contribute to the complexity of the scenario (Ignat et al., 2011). In addition, the chemical composition of foods is strongly influenced by countless factors, such as the type of cultivar, the growing conditions, and the seasonality, among others (Dias et al., 2021). This phytochemical concentration variability has been deeply considered in this work by collecting multiple data sources, and it represents a critical point when assessing the dietary intake of plant bioactives in a population. Therefore, further efforts to enhance the quality of the data could be implemented by adding supplementary information about the food, such as the type of cultivars and different processing techniques used. Finally, we reported more than 60% of the PhytoFooD data show a robustness index of 1; this implies that, for each phytochemical, up to three concentration data have been used to obtain the final value reported in our database, underlining the scarcity of information currently available. However, inclusion and exclusion criteria were set during the literature screening to ensure a high quality of the information collected, which has favoured data quality over data quantity.

To provide insights into the PhytoFooD potential applications, we estimated the intake of major phytochemicals using average consumption data of the 20 most consumed food items in the European adult’s diet. However, these results should be considered as representative examples of PhytoFooD uses -rather than a comprehensive picture of the dietary exposure of plant bioactives-, since a limited number of food items and phytochemicals were considered. Further actions will be pursued along this path, in terms of completeness of phytochemicals and foods investigated, to provide exhaustive and referential information on the dietary phytochemical intake in different populations. Finally, this work represents a pioneering effort to develop a comprehensive phytochemical food composition database, which will be used to assess the intake of plant bioactives in multiple populations.

## Data Availability

All data produced in the present study are available upon reasonable request to the authors

## Acknowledgements

This work was supported by the European Research Council (ERC) under the European Union’s Horizon 2020 research and innovation programme (PREDICT-CARE project, grant agreement No 950050) and it has received funding from the Italian Ministry for Universities and Research (MUR) under the FARE programme (CARE-DIET, R20MPBW4FM). This work has been carried out in the frame of the ALIFAR project, funded by the Italian Ministry of University through the program ‘Dipartimenti di Eccellenza 2023-2027’. Authors thank the students that participated in data collection and harmonization, and the colleagues of Human Nutrition Unit of the University of Parma for providing support on literature search and food analysis techniques.

## Author contributions

C.Mic., A.R., F.S., D.D.R, and P.M. conceptualized the study. C.Mic., A.R. and F.B. collected and assembled the composition data, and developed the model code for the analysis. C.Mig. performed the clinical data and collected dietary data. C.Mic. and F.B. analysed the data. C.Mic. drafted the paper. P.M. obtained funding for the study. All authors interpreted the results, revised the paper for important intellectual content, and read and approved the final paper.

## Supplementary materials

**Table S1:**
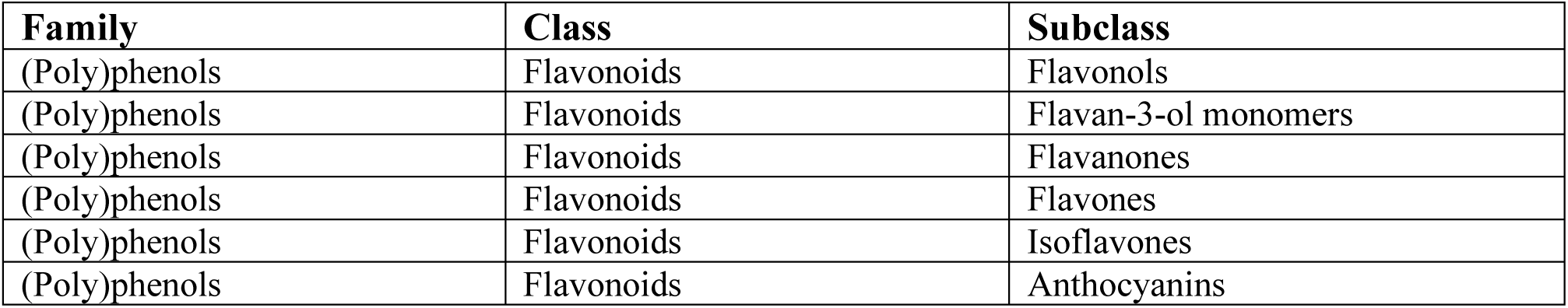
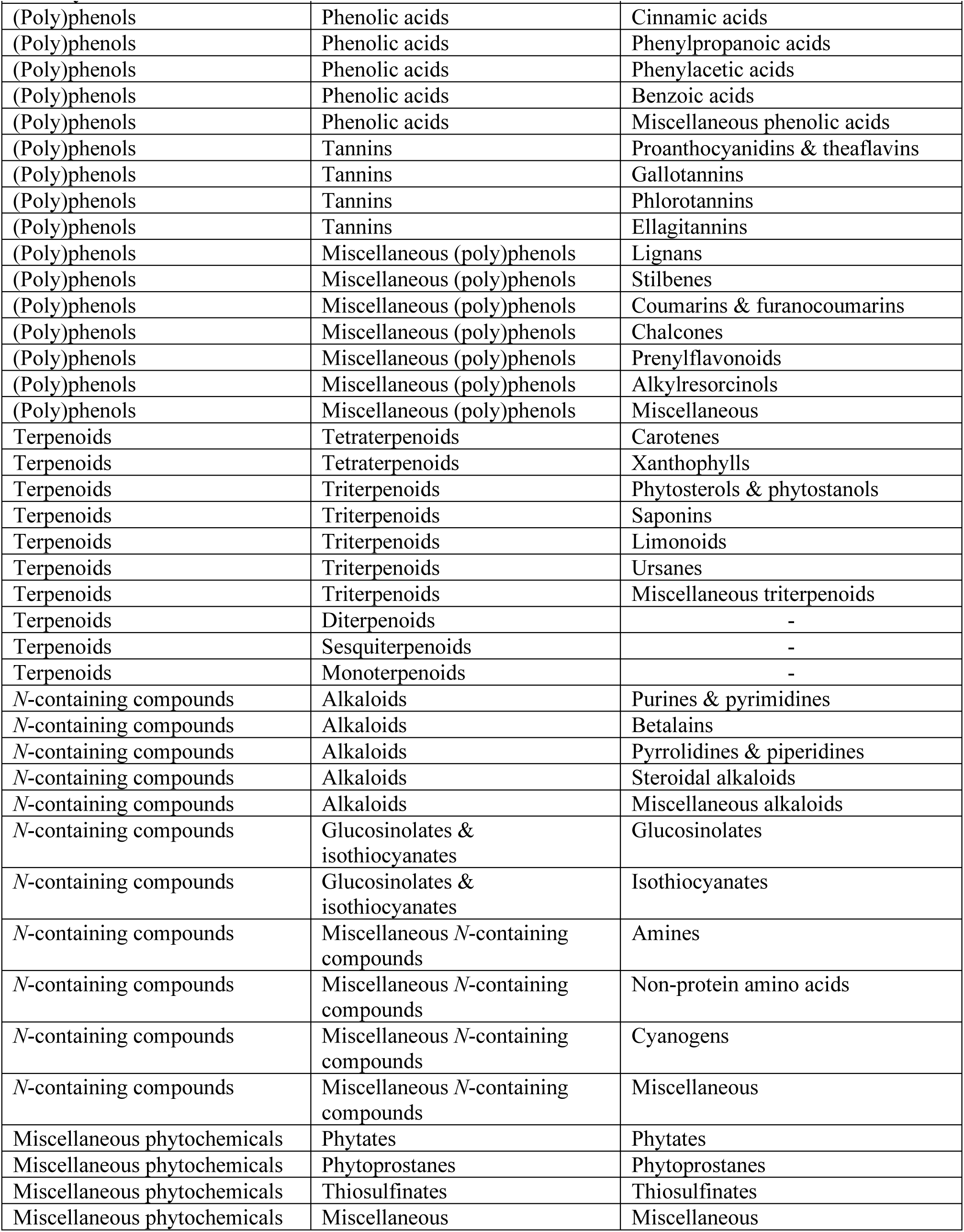
Dietary phytochemical classification. Plant-based foods represent an enormous storage of bioactive molecules with incredibly different molecular structures. This classification combines PhytoHub classification and current trends from the literature.

**Figure S1:**
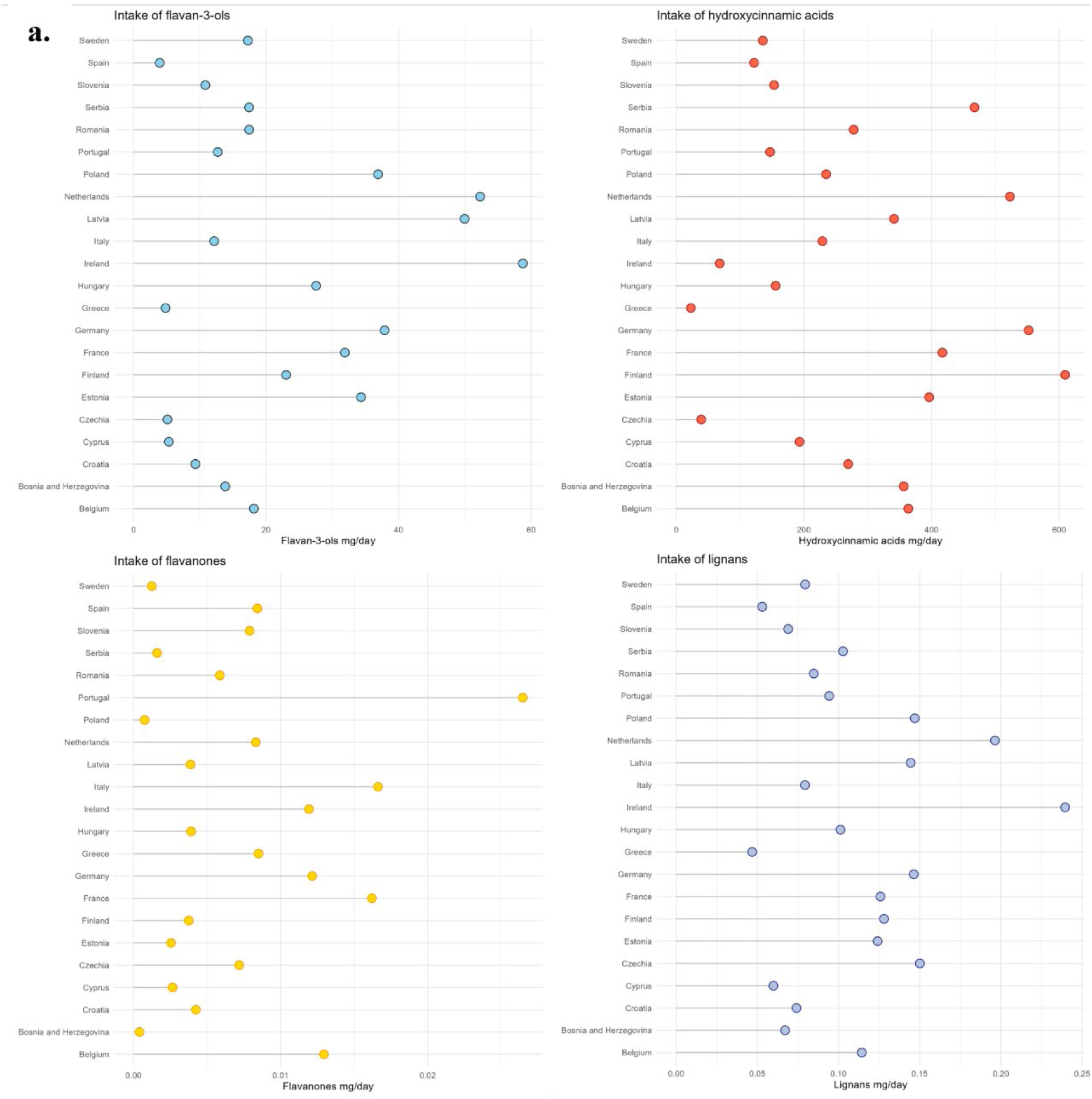
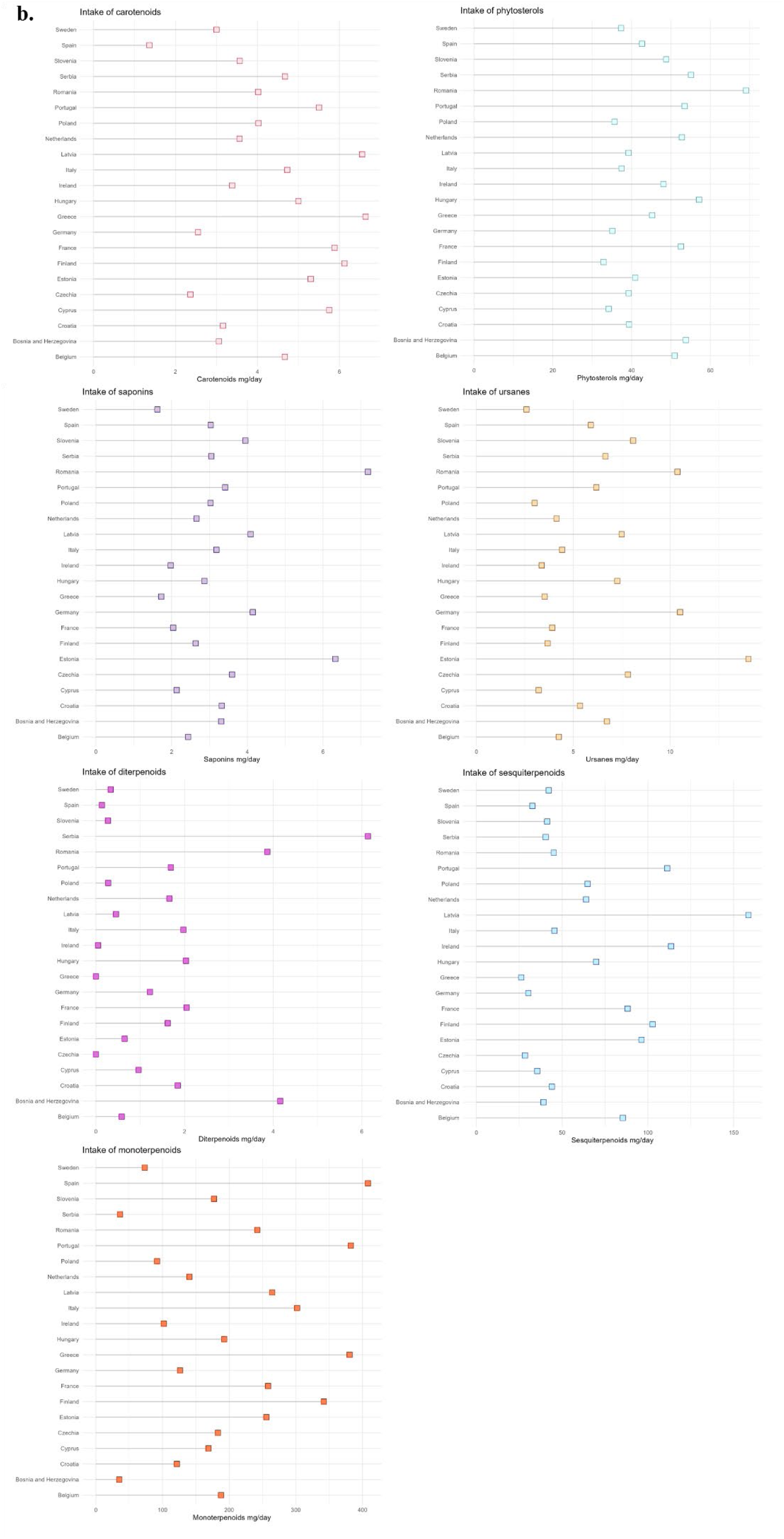
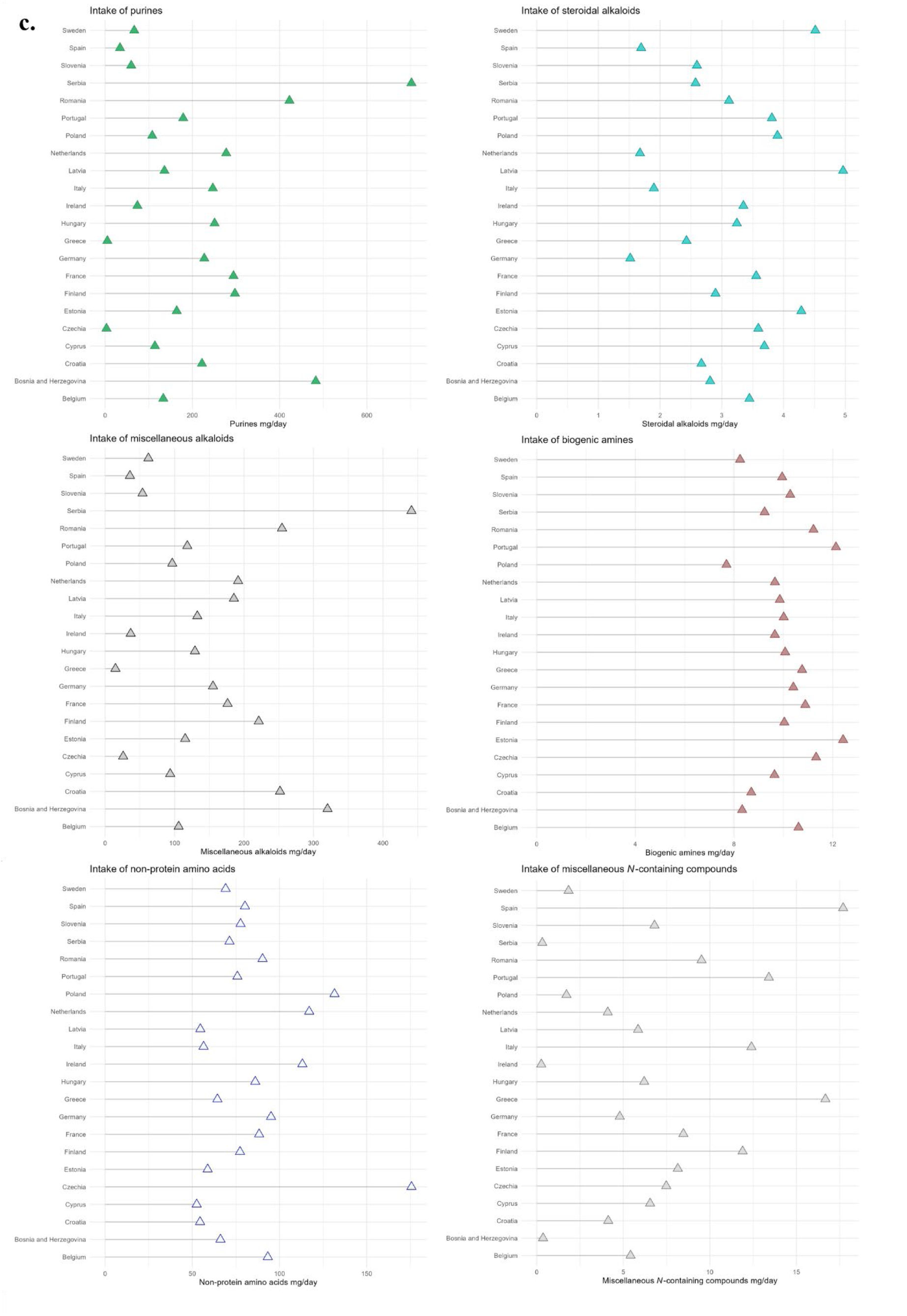
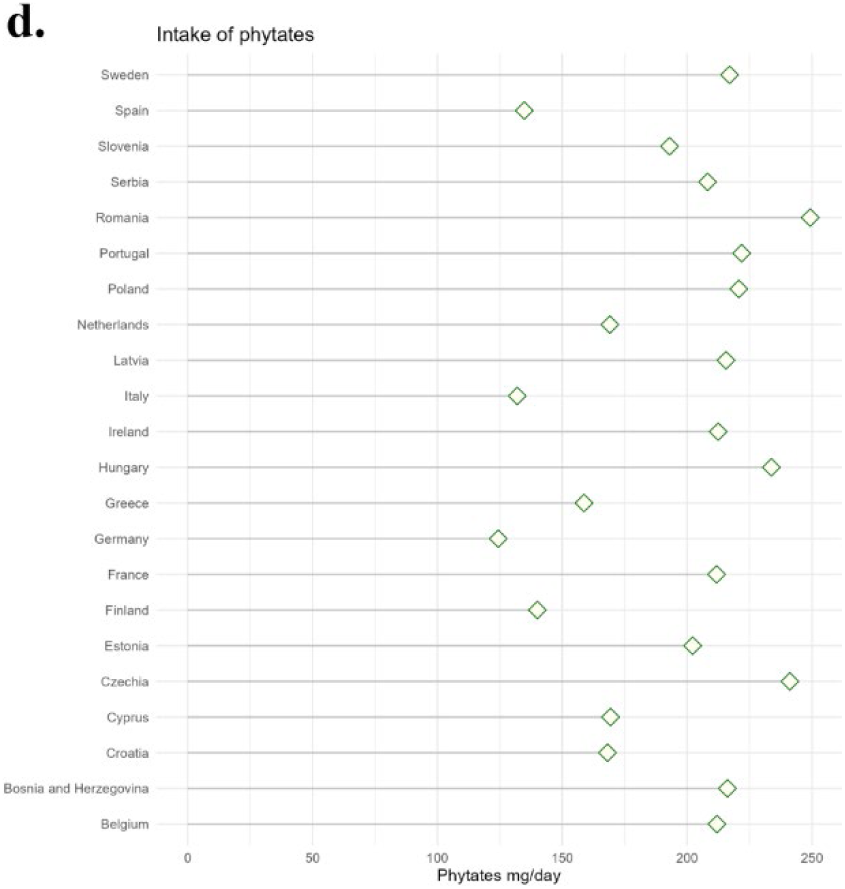
Intake of phytochemical classes and subclasses in the European population. The intake of selected phytochemicals from the 20 most consumed food items from the European populations have been estimated. Here are represented the intake of plant bioactives, gathered in classes and subclasses of (poly)phenols (**a**), terpenoids (**b**), *N*-containing compounds (**c**) and miscellaneous phytochemicals (**d**).

