## Supplementary material for "Development and application of a Phytochemical Food Database (PhytoFooD) to assess the intake of dietary plant bioactives": Table S1 & Figure S1

**Table S1: Dietary phytochemical classification.** Plant-based foods represent an enormous storage of bioactive molecules with incredibly different molecular structures. This classification combines PhytoHub classification and current trends from the literature.

| **Family** | **Class** | **Subclass** |
|---|---|---|
| (Poly)phenols | Flavonoids | Flavonols |
| (Poly)phenols | Flavonoids | Flavan-3-ol monomers |
| (Poly)phenols | Flavonoids | Flavanones |
| (Poly)phenols | Flavonoids | Flavones |
| (Poly)phenols | Flavonoids | Isoflavones |
| (Poly)phenols | Flavonoids | Anthocyanins |
| (Poly)phenols | Phenolic acids | Cinnamic acids |
| (Poly)phenols | Phenolic acids | Phenylpropanoic acids |
| (Poly)phenols | Phenolic acids | Phenylacetic acids |
| (Poly)phenols | Phenolic acids | Benzoic acids |
| (Poly)phenols | Phenolic acids | Miscellaneous phenolic acids |
| (Poly)phenols | Tannins | Proanthocyanidins & theaflavins |
| (Poly)phenols | Tannins | Gallotannins |
| (Poly)phenols | Tannins | Phlorotannins |
| (Poly)phenols | Tannins | Ellagitannins |
| (Poly)phenols | Miscellaneous (poly)phenols | Lignans |
| (Poly)phenols | Miscellaneous (poly)phenols | Stilbenes |
| (Poly)phenols | Miscellaneous (poly)phenols | Coumarins & furanocoumarins |
| (Poly)phenols | Miscellaneous (poly)phenols | Chalcones |
| (Poly)phenols | Miscellaneous (poly)phenols | Prenylflavonoids |
| (Poly)phenols | Miscellaneous (poly)phenols | Alkylresorcinols |
| (Poly)phenols | Miscellaneous (poly)phenols | Miscellaneous |
| Terpenoids | Tetraterpenoids | Carotenes |
| Terpenoids | Tetraterpenoids | Xanthophylls |
| Terpenoids | Triterpenoids | Phytosterols & phytostanols |
| Terpenoids | Triterpenoids | Saponins |
| Terpenoids | Triterpenoids | Limonoids |
| Terpenoids | Triterpenoids | Ursanes |
| Terpenoids | Triterpenoids | Miscellaneous triterpenoids |
| Terpenoids | Diterpenoids | - |
| Terpenoids | Sesquiterpenoids | - |
| Terpenoids | Monoterpenoids | - |
| *N*-containing compounds | Alkaloids | Purines & pyrimidines |
| *N*-containing compounds | Alkaloids | Betalains |
| *N*-containing compounds | Alkaloids | Pyrrolidines & piperidines |
| *N*-containing compounds | Alkaloids | Steroidal alkaloids |
| *N*-containing compounds | Alkaloids | Miscellaneous alkaloids |
| *N*-containing compounds | Glucosinolates & isothiocyanates | Glucosinolates |
| *N*-containing compounds | Glucosinolates & isothiocyanates | Isothiocyanates |
| *N*-containing compounds | Miscellaneous *N*-containing compounds | Amines |
| *N*-containing compounds | Miscellaneous *N*-containing compounds | Non-protein amino acids |
| *N*-containing compounds | Miscellaneous *N*-containing compounds | Cyanogens |

| Family | Class | Subclass |
|---|---|---|
| *N*-containing compounds | Miscellaneous *N*-containing compounds | Miscellaneous |
| Miscellaneous phytochemicals | Phytates | Phytates |
| Miscellaneous phytochemicals | Phytoprostanes | Phytoprostanes |
| Miscellaneous phytochemicals | Thiosulfinates | Thiosulfinates |
| Miscellaneous phytochemicals | Miscellaneous | Miscellaneous |

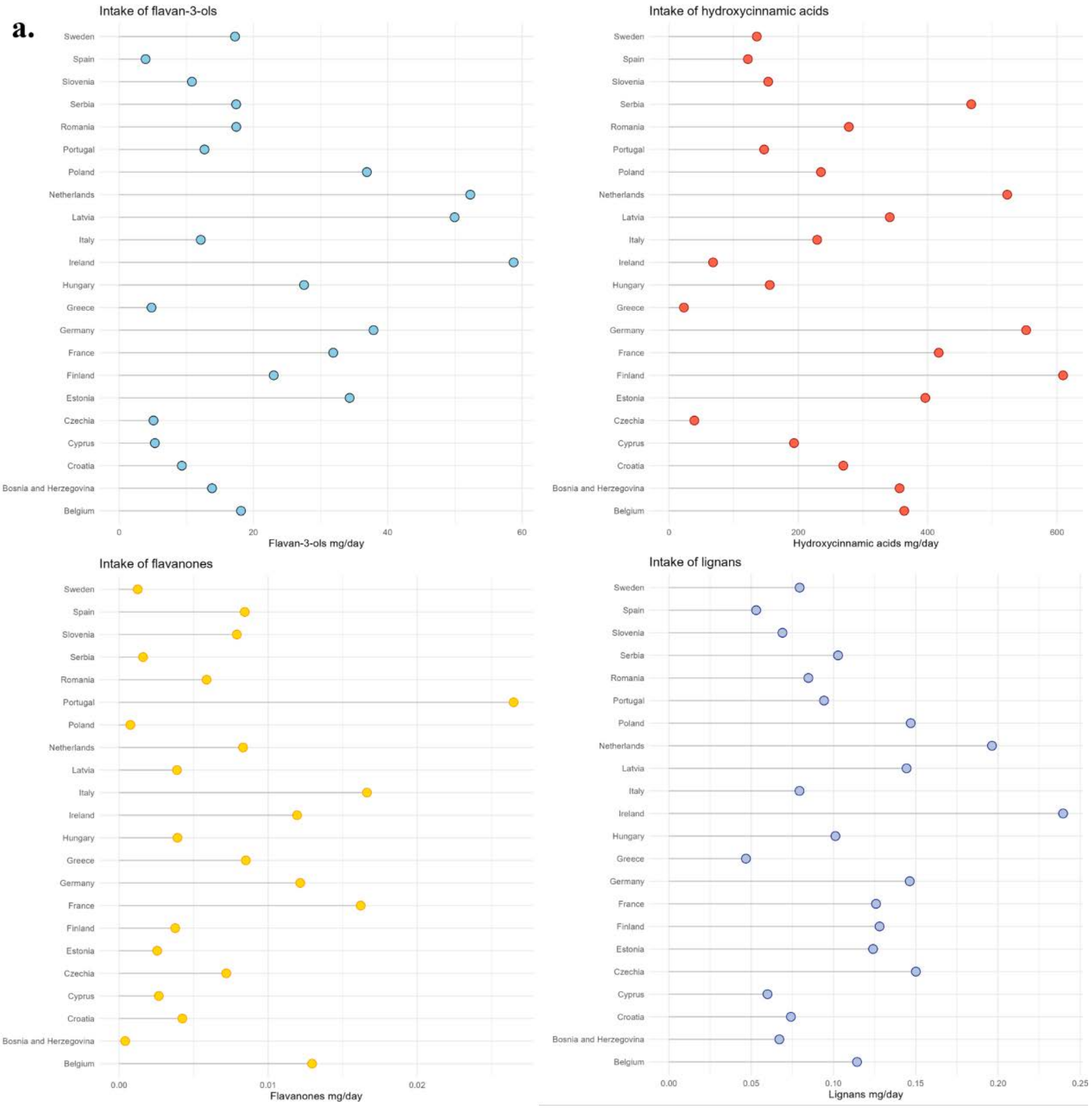

**b.**

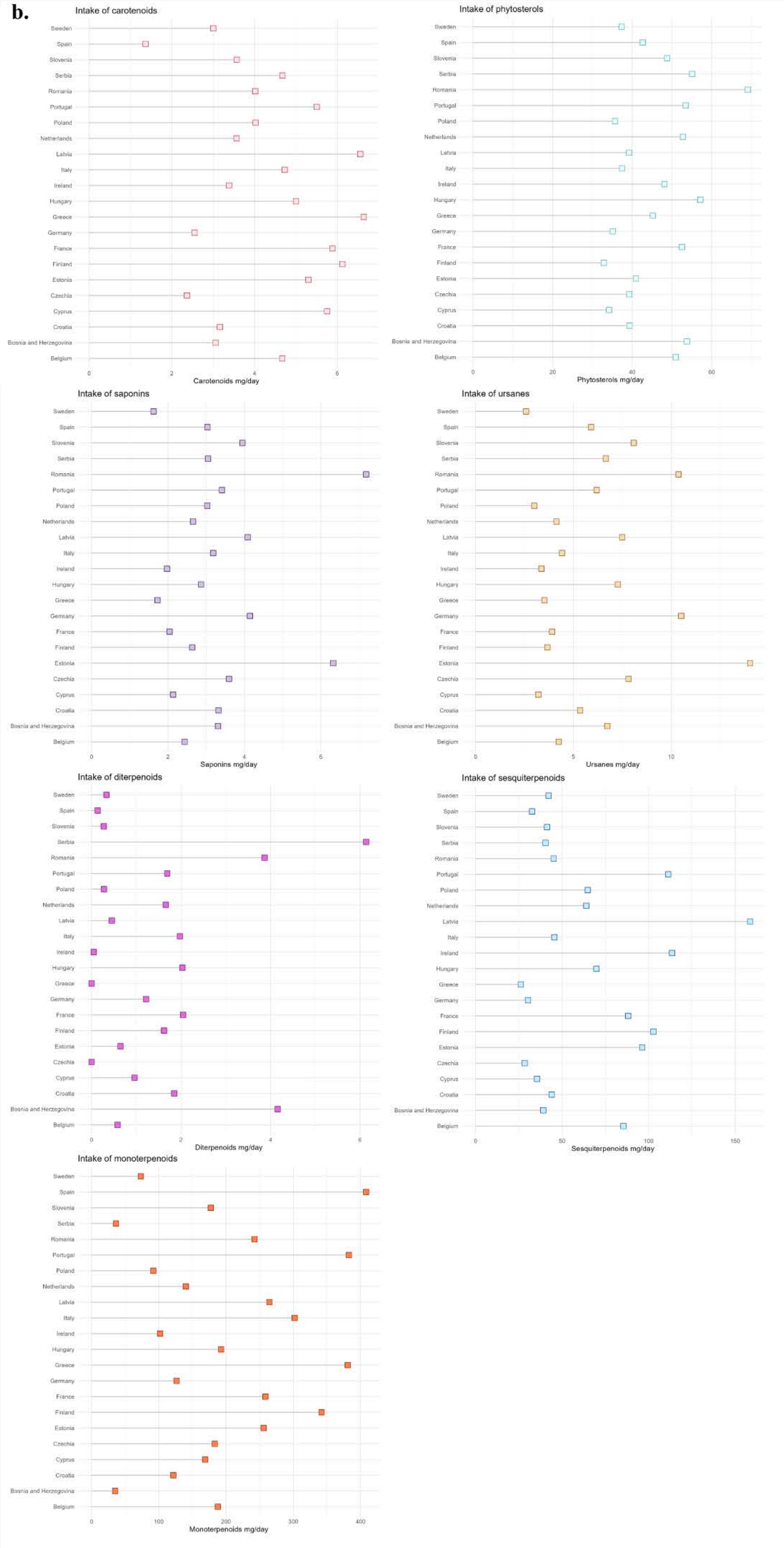
Intake of carotenoids
Sweden
Spain
Slovenia
Serbia
Romania
Portugal
Poland
Netherlands
Latvia
Italy
Ireland
Hungary
Greece
Germany
France
Finland
Estonia
Czechia
Cyprus
Croatia
Bosnia and Herzegovina
Belgium
0
2
4
6
Carotenoids mg/day
Intake of phytosterols
0
20
40
60
Phytosterols mg/day
Intake of saponins
0
2
4
6
Saponins mg/day
Intake of ursanes
0
5
10
Ursanes mg/day
Intake of diterpenoids
0
2
4
6
Diterpenoids mg/day
Intake of sesquiterpenoids
0
50
100
150
Sesquiterpenoids mg/day
Intake of monoterpenoids
0
100
200
300
400
Monoterpenoids mg/day

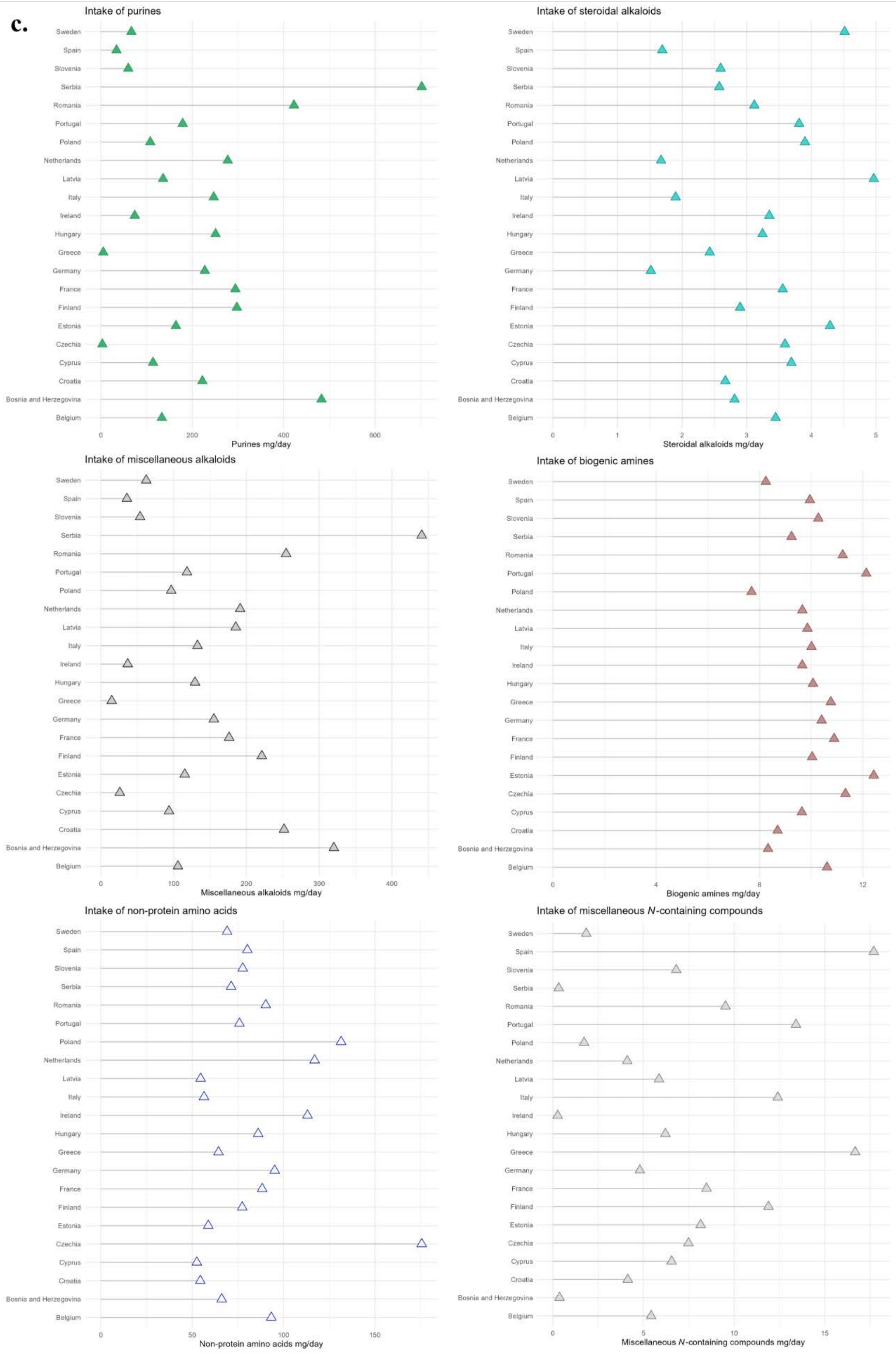

c.
Intake of purines
Purines mg/day
Intake of steroidal alkaloids
Steroidal alkaloids mg/day
Intake of miscellaneous alkaloids
Miscellaneous alkaloids mg/day
Intake of biogenic amines
Biogenic amines mg/day
Intake of non-protein amino acids
Non-protein amino acids mg/day
Intake of miscellaneous N-containing compounds
Miscellaneous N-containing compounds mg/day
Sweden
Spain
Slovenia
Serbia
Romania
Portugal
Poland
Netherlands
Latvia
Italy
Ireland
Hungary
Greece
Germany
France
Finland
Estonia
Czechia
Cyprus
Croatia
Bosnia and Herzegovina
Belgium

**d.**

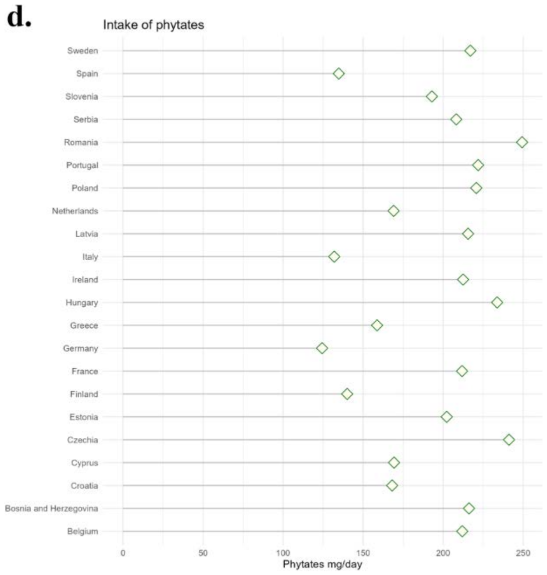


**Figure S1: Intake of phytochemical classes and subclasses in the European population.** The intake of selected phytochemicals from the 20 most consumed food items from the European populations have been estimated. Here are represented the intake of plant bioactives, gathered in classes and subclasses of (poly)phenols (**a**), terpenoids (**b**), *N*-containing compounds (**c**) and miscellaneous phytochemicals (**d**).
